# Sex-specific trends in incident stroke: The Framingham Heart Study

**DOI:** 10.64898/2026.04.22.26351536

**Authors:** Oluchi Ekenze, Matthew Scott, Dibya Himali, Vasileios-Arsenios Lioutas, Sudha Seshadri, Virginia J. Howard, Myriam Fornage, Hugo J. Aparicio, Alexa S. Beiser, Jose R. Romero

## Abstract

Sex specific differences in stroke are recognized. Whether differences in incident stroke risk persists in recent periods needs further elucidation to aid public health preventive efforts.

**Aim:** To determine long-term sex specific trends in stroke and stroke risk factors at different epochs among Framingham Heart Study participants.

**Methods:** We examined age-adjusted 10-year stroke incidence using Cox regression in women and men in five epochs: 1962-1969 (epoch 1, reference), 1971-1976 (epoch 2), 1987-1991 (epoch 3), 1998-2005 (epoch 4), 2015-2021 (epoch 5). We compared stroke incidence by sex across epochs, estimated decade-wise linear trends overall and by sex. We compared risk factors in successive epochs to the first, and estimated sex-specific trends in risk factors. Interactions between baseline risk factors with epoch and trends were assessed by sex. Secondary analyses were repeated in participants <60 years old.

**Results:** Incident stroke occurred in 4.5% (178/3996) in epoch 1, 3.9% (227/5786) in epoch 2, 3.9% (199/5137) in epoch 3, 2.7% (207/7642) in epoch 4, 2.2% (119/5534) in epoch 5. Men had higher risk of incident stroke in each epoch with significant difference in epochs 2 (HR 1.41, 95% CI [1.08, 1.84]) and 4 (HR 1.46, 95% CI [1.11, 1.91]) overall, and in epoch 4 (HR 2.13, 95% CI [1.17, 3.87]) among those <60 years. Stroke incidence declined by 16% per decade in men (HR 0.84, 95% CI [0.79, 0.89]) and 19% per decade in women (HR 0.81, 95% CI [0.76, 0.86]). Among those <60 years, stroke incidence declined by 22% per decade in women (HR 0.78, 95% CI [0.67, 0.95]). Hypertension declined by 8% per decade in women only ([OR] 0.92, 95% CI [0.90, 0.94]), while Atrial fibrillation and diabetes increased in both.

**Conclusion:** Stroke incidence continues to decline in recent periods for women and men. Among participants <60 years, decline was observed only in women, possibly related to decline in hypertension in women.

## Background

Stroke remains a major contributor to the global burden of disease, and a foremost cause of death and disability-adjusted life years worldwide.^1^ Globally, the incidence of stroke is projected to increase by 81% from 11.81million in 2021 to 21.43million in 2050.^2^ In the US, about 9million adults > 20 years have a stroke.^3^ Although men have higher stroke incidence and global burden of stroke,^2,4^ stroke incidence in women is predicted to exceed men by 2050.^2^

Stroke incidence varies between men and women across the life span. For example, in the US REasons for Geographic and Racial Differences in Stroke (REGARDS) study, both Black and White women aged 45 to 64 had a lower stroke risk than men, but there was no difference by sex among those aged 75 and older.^5^ Prior reports from the Framingham Heart Study (FHS), showed stroke incidence was higher in men between the ages of 45 and 84years, while at ages 85 to 94years it was higher in women.^6^ Among community dwellers in Ontario Canada, compared to men, women had higher stroke incidence among those less than 30 years, lower incidence in those aged 40 years - 80 years and similar incidence as men in those 80 years and older.^1^

Temporal trends in stroke incidence by sex have been inconsistent across studies. Some studies report a greater decline in stroke incidence over time in men,^7,8^ others in women,^9,10^ whereas others detect no significant sex differences.^11–15^ Further in the Greater Cincinnati Kentucky Stroke Study, trends in stroke incidence were age-specific with higher incidence in men aged 20-44 years, decline in stroke incidence in both sexes in those aged 65-84 years and more decline in men aged 85 years and older.^15^ The lifetime risk for stroke is higher in women than men.^16,17^

Sex differences in stroke trends may reflect differential risk factor burden. Although women and men share major stroke risk factors, their relative impact differs. For instance, in the INTERSTROKE study, hypertension, waist-to-hip ratio and dyslipidemia were the most impactful risk factors for stroke in women whereas smoking and cardiac disease were infrequent.^18^ In the REasons for Geographic And Racial Differences in Stroke (REGARDS) study cohort, the severity of hypertension was associated with twice the risk of ischemic stroke in women compared to men.^19^ Further, among the UK Biobank cohort, there was a stronger association between hypertension and risk of incident stroke in women compared to men.^20^ The association between atrial fibrillation and stroke has been inconsistent. Some studies suggest that women with atrial fibrillation have a higher risk of stroke,^20–26^ while others report no association particularly in those <75 years.^27^ Further, studies have shown abdominal obesity to be significantly associated with stroke in women but not men, ^28–30^ women with diabetes having higher stroke risk compared to men,^20,31–34^ and smoking associated with higher stroke risk in men. Additionally, with advancing age, women may experience greater increase in blood pressure, total cholesterol levels compared to men.^3^

Although prior studies have observed stroke incidence has decreased overtime,^25^ analyses that explicitly assess sex-specific secular trends remain limited. Clarifying whether declines in stroke incidence have occurred similarly in men and women is critical to developing health policies and preventive strategies to reduce stroke burden among highest risk groups. Additionally, sex-specific trends in stroke among younger individuals remain underexplored. To address this knowledge gap, we examined sex-specific trends in stroke incidence and investigated potential determinants of observed sex differences using longitudinal data from the FHS.

## METHODS

### Study Sample

The FHS has been previously described.^35^ The FHS started in 1948 with the recruitment of the original cohort. This was followed by recruitment of the offspring of the original cohort and spouses of the offspring in 1971. Subsequently, the third generation (Gen 3) including grandchildren of the original cohort was recruited in 2002. To add to the familial data, parents of the Gen 3 participants who were not previously enrolled in the Offspring cohort were enrolled in 2003 as the New Offspring Spouse cohort (NOS). The Omni 1 cohort representing more ethnic and racial diversity of the town of Framingham was recruited in 1994. To expand from the Omni Cohort 1 and to represent an ethnically diverse group at least 10% of the size of the Gen 3 cohort, enrollment of Omni 2 cohort started in 2003. Omni Cohort 2 included individuals related and unrelated to the participants of Omni Cohort 1.

Longitudinal, systematic examination of FHS participants has been done over seven decades. The Original cohort was examined every 2 years, the Offspring and Omni 1 cohorts every 4-8 years, the Gen 3 & Omni 2 cohorts every 4-8 years. Comprehensive clinical assessments have been done including detailed physical examination, anthropometric measurements, cardiovascular and neurological assessments, collection of blood and urine for laboratory examinations and participation in on-site ancillary studies.^35^ Between exam cycles, interim surveys detailing updates of medical and family history are mailed, and information is obtained via regular phone calls and electronic mails, thereby maintaining continuous surveillance for incident stroke in all the participants. The 99% retention rate of participants regularly returning for scheduled examinations at FHS is a demonstration of the commitment of participants and researchers to the study and impacts to the high quality of the study.^35^

For the present study, we included participants from all cohorts who attended examinations in any of 5 epochs: epoch 1: 1962-1967 (original cohort exam 8), epoch 2: 1971-1976 (original cohort exam 13 and offspring cohort exam 1), epoch 3: 1987-1991 (original cohort exam 20, offspring cohort exam 4), epoch 4: 1998-2005 (original cohort exam 26, offspring cohort exam7, Omni 1 cohort exam 2, and Gen 3 and Omni 2 cohorts exam 1), and epoch 5: 2015-2021 (offspring cohort exam 9, Omni 1 cohort exam 4, and Gen 3 and Omni 2 cohorts exam 3). We chose these different epochs *a priori* as they coincided with diagnostic or treatment advancements in stroke, such as the availability of CT scan or MRI for the diagnosis of stroke or more recent changes in standard of care as mechanical thrombectomy procedures were incorporated in clinical practice, and prior FHS research has shown a difference in the burden of stroke risk factors during these epochs.^36^.

### Outcome

Clinical stroke was defined as rapidly developing signs of focal neurologic disturbance of presumed vascular etiology lasting more than 24 hours. This definition excludes transient ischemic attacks and silent cerebral infarctions or hemorrhages detected only by imaging.^37^

### Stroke determination

Surveillance for incident stroke events involves continuous daily monitoring of participant admissions to local hospitals and emergency room visits in Framingham and other areas, following records for interim hospitalizations, enquiring for stroke symptoms in annual telephone health updates, screening at consecutive cohort examinations, referrals from ancillary study examinations, primary care providers, study participants or their family members.

Participants with suspected stroke undergo a standardized evaluation and final determination of stroke and stroke subtype is done by a panel of three FHS stroke neurologists. The diagnosis of stroke is based on preestablished criteria that include review of all available data including medical records, acute stroke treatment, and available imaging studies (CT/MRI), vascular studies (CTA, MRA and conventional angiography, carotid ultrasound), cardiac evaluations (electrocardiogram, trans-thoracic or trans-esophageal echocardiography, cardiac event monitor, implantable loop recorder), and when available information from autopsy studies among participants in the FHS brain donation program.^37^

The study was approved by the Institutional Review board of Boston University Medical Center.

#### Covariates

Baseline demographic and clinical characteristics for the participants were obtained at the start of the follow-up in each epoch. Prevalent diabetes is defined as random blood glucose ≥ 200 mg/dl (11.1 mmol/L) or fasting blood sugar ≥ 126 mg/dL (7 mmol/L) or the use of oral hypoglycemic agents or insulin for all cohorts. Hypertension, according to Seventh Report of the Joint National Committee on Prevention, Detection, Evaluation, and Treatment of High Blood Pressure Criteria, was defined as a systolic blood pressure of ≥140 mmHg, a diastolic blood pressure of ≥90 mmHg, or the use of antihypertensive medications.^38^ Current smoking status was self-reported smoking of at least 1 cigarette per day within the year preceding examination. The use of preventive medications was self-reported. Body mass index (BMI) was defined as weight (Kg) divided by height (m^2^). Total cholesterol (mg/dL) was measured on fasting specimens.

Prevalent cardiovascular disease was defined as presence of coronary heart disease (stable angina, coronary insufficiency, and myocardial infarction) or heart failure. These diagnoses were adjudicated according to the established FHS criteria by a panel of three physicians from endpoint review.^35,39^ Atrial fibrillation was diagnosed if at least two FHS cardiologists confirmed rhythm abnormality based on ECG, including external Holter ECGs, telemetry, or other monitoring data.^39^ Cholesterol-lowering medications use was self-reported and confirmed by review of medications brought to each follow-up visit.

### Statistical Analysis

The sex-specific, epoch-specific baseline demographic and clinical characteristics were obtained at the start of follow-up in each epoch and described using mean (SD) for continuous variables and count (%) for categorical variables. To determine trends in vascular risk factors, we examined sex-specific trends in risk factors during all epochs compared to epoch 1 as reference by regressing the baseline characteristics on median time from baseline within each epoch, adjusting for age. Further, we assessed the interaction of sex and epoch/trends in their association with baseline characteristics.

We used Cox proportional-hazard models to estimate 10-year hazard ratios (HR) for incident stroke during each epoch, adjusting for age at baseline of each epoch. To account for repeated measures across epochs, Sandwich estimators^40^ were applied to all Cox models. Participants without stroke were censored at death or up to 10 years after their epoch-specific baseline clinic exam.

Within each epoch, we estimated the HR for incident stroke in men compared to women. Further, we estimated the HR for incident stroke in successive epochs relative to the first epoch and estimated linear trends (the decline per decade in 10-year incidence of stroke) using elapsed median time in decades between the first epoch and each successive epoch. We assessed interactions with sex in these analyses.

In secondary analysis, to determine any age differences in stroke incidence over time, we repeated the above analyses restricting the sample to participants aged < 60 years. We chose age 60 years cut off because of lack of the protective effect of endogenous sex steroids among older women.^41^ Due to estimation issues relating to low event counts, temporal trends using Firth correction in participants < 60 years are reported in the Supplement.

A threshold of p value <0.1 was used to identify significant interaction. Data analysis was performed using SAS version 9.4. For all other analyses, we used a two-sided threshold of p<0.05

## Results

Women and men aged ≥ 35 years without prevalent stroke contributed to 15,491 and 12,574 observations respectively.

### Trends in baseline characteristics

Sex-stratified analyses of trends in risk factors over time revealed similar and differing trends depending on individual risk factors. In both sexes, there was significant decrease in trends for prevalent smoking, prevalent CVD, total cholesterol, and total cholesterol/ HDL ratio over time while the trends for prevalent atrial fibrillation, prevalent diabetes, and treatment for diabetes and hypertension increased over time. The trends for prevalence of hypertension decreased over time only in women. (Tables 1A &1B). There was a multiplicative interaction of sex and epoch/trend with baseline characteristics such that over time, women were more likely to smoke, had less hypertension and had higher Total cholesterol/HDL ratio than men while men over time had more diabetes than women. (Table 2).

**Table 1:**
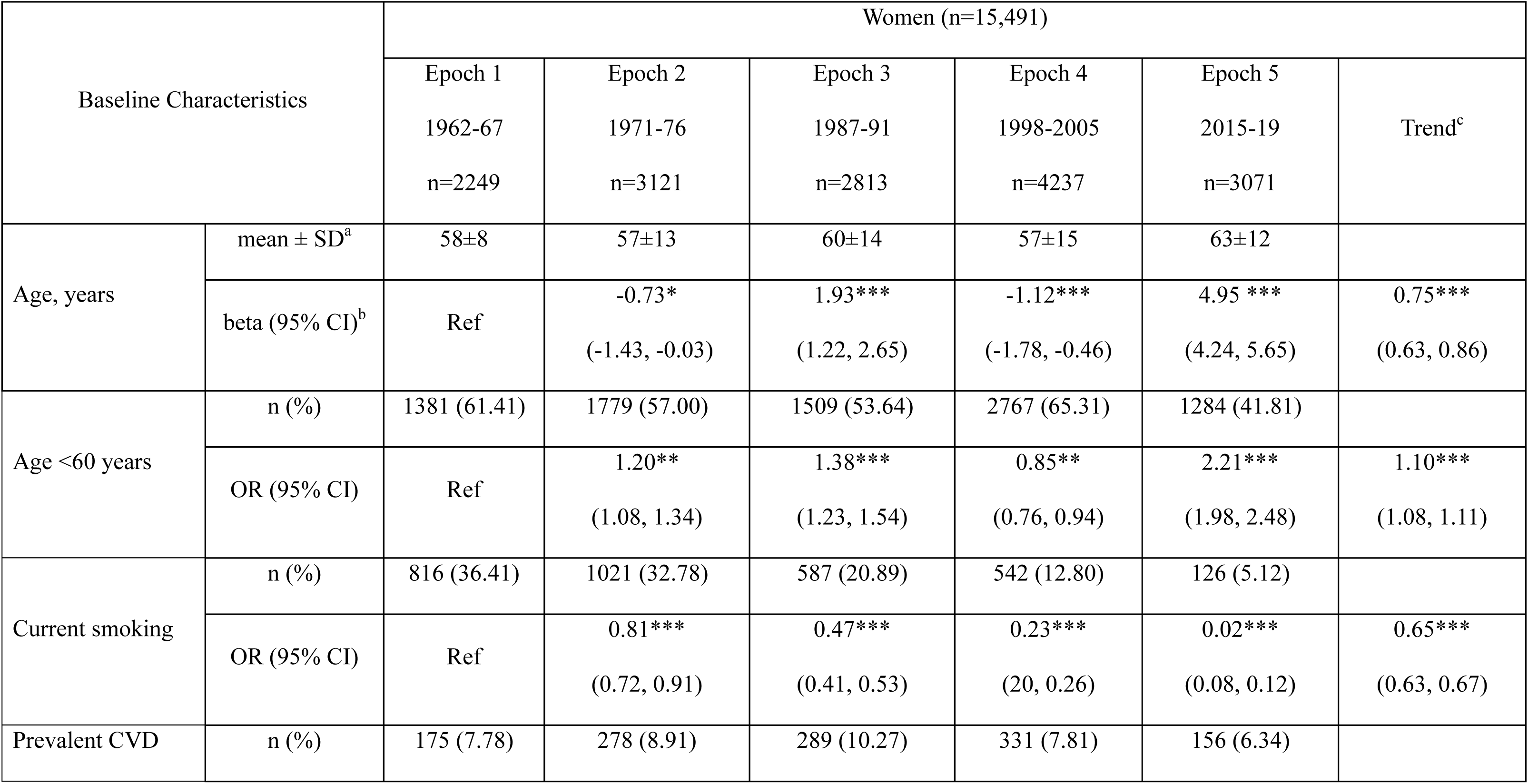
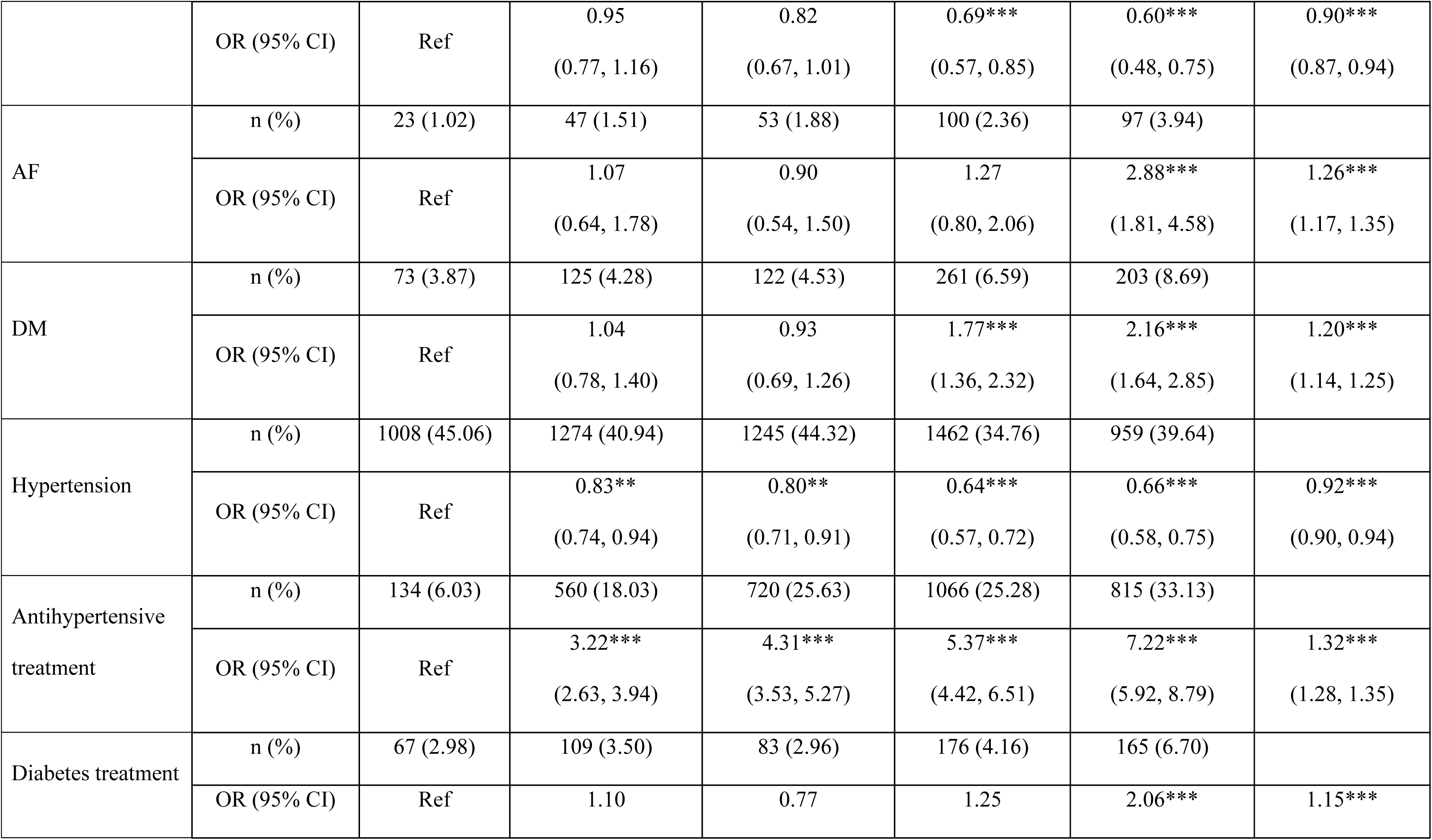
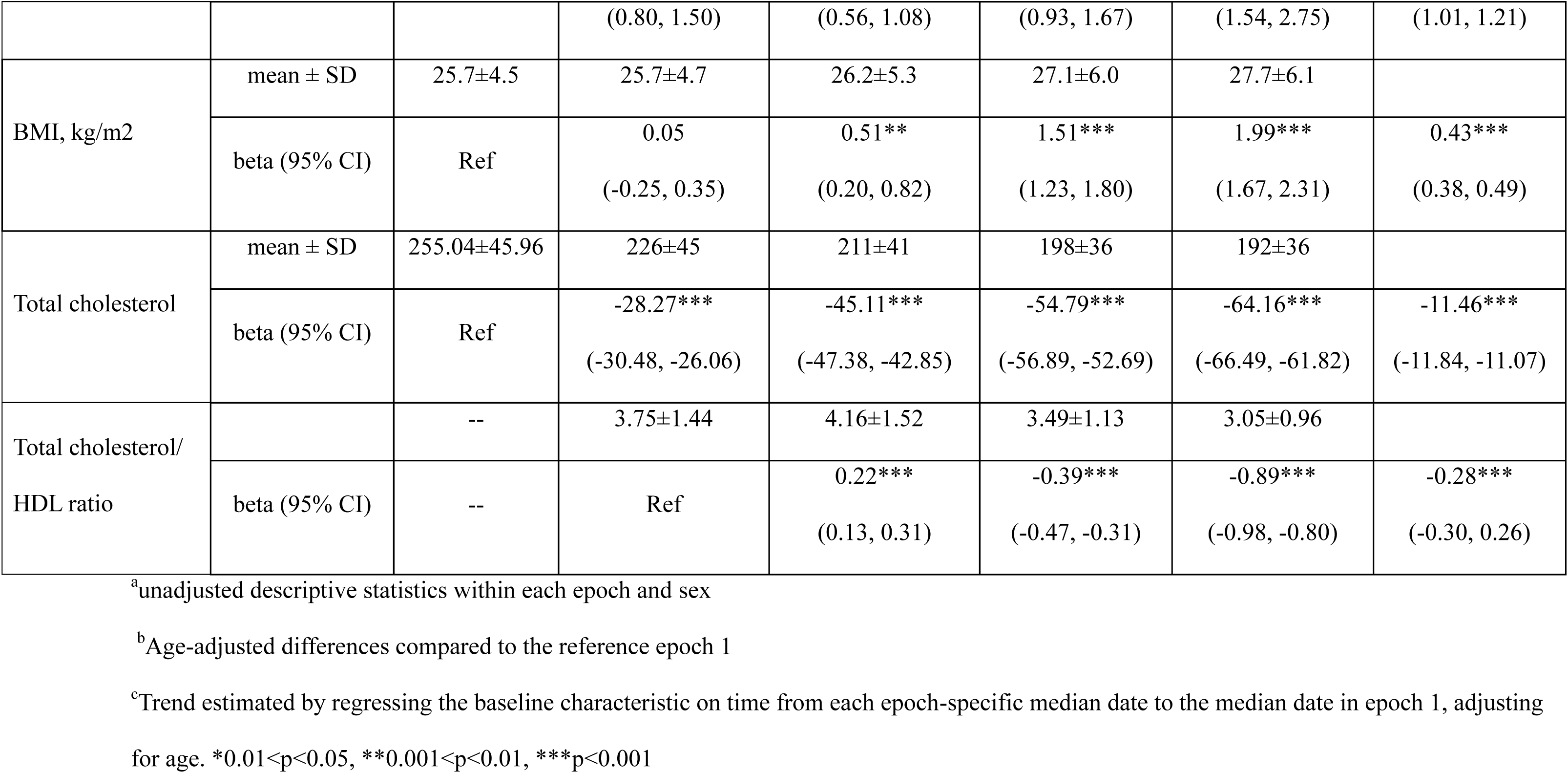
Baseline Characteristics and Incident Stroke Events in Each Epoch, in women.

| Baseline Characteristics |  | Women (n=15,491) |  |  |  |  |  |
| --- | --- | --- | --- | --- | --- | --- | --- |
|  |  | Epoch 1<br>1962-67<br>n=2249 | Epoch 2<br>1971-76<br>n=3121 | Epoch 3<br>1987-91<br>n=2813 | Epoch 4<br>1998-2005<br>n=4237 | Epoch 5<br>2015-19<br>n=3071 | Trend<sup>c</sup> |
| Age, years | mean ± SD<sup>a</sup> | 58±8 | 57±13 | 60±14 | 57±15 | 63±12 |  |
|  | beta (95% CI)<sup>b</sup> | Ref | -0.73*<br>(-1.43, -0.03) | 1.93***<br>(1.22, 2.65) | -1.12***<br>(-1.78, -0.46) | 4.95 ***<br>(4.24, 5.65) | 0.75***<br>(0.63, 0.86) |
| Age <60 years | n (%) | 1381 (61.41) | 1779 (57.00) | 1509 (53.64) | 2767 (65.31) | 1284 (41.81) |  |
|  | OR (95% CI) | Ref | 1.20**<br>(1.08, 1.34) | 1.38***<br>(1.23, 1.54) | 0.85**<br>(0.76, 0.94) | 2.21***<br>(1.98, 2.48) | 1.10***<br>(1.08, 1.11) |
| Current smoking | n (%) | 816 (36.41) | 1021 (32.78) | 587 (20.89) | 542 (12.80) | 126 (5.12) |  |
|  | OR (95% CI) | Ref | 0.81***<br>(0.72, 0.91) | 0.47***<br>(0.41, 0.53) | 0.23***<br>(0.20, 0.26) | 0.02***<br>(0.08, 0.12) | 0.65***<br>(0.63, 0.67) |
| Prevalent CVD | n (%) | 175 (7.78) | 278 (8.91) | 289 (10.27) | 331 (7.81) | 156 (6.34) |  |
|  | OR (95% CI) | Ref | 0.95<br>(0.77, 1.16) | 0.82<br>(0.67, 1.01) | 0.69***<br>(0.57, 0.85) | 0.60***<br>(0.48, 0.75) | 0.90***<br>(0.87, 0.94) |
| AF | n (%) | 23 (1.02) | 47 (1.51) | 53 (1.88) | 100 (2.36) | 97 (3.94) |  |
|  | OR (95% CI) | Ref | 1.07<br>(0.64, 1.78) | 0.90<br>(0.54, 1.50) | 1.27<br>(0.80, 2.06) | 2.88***<br>(1.81, 4.58) | 1.26***<br>(1.17, 1.35) |
| DM | n (%) | 73 (3.87) | 125 (4.28) | 122 (4.53) | 261 (6.59) | 203 (8.69) |  |
|  | OR (95% CI) | Ref | 1.04<br>(0.78, 1.40) | 0.93<br>(0.69, 1.26) | 1.77***<br>(1.36, 2.32) | 2.16***<br>(1.64, 2.85) | 1.20***<br>(1.14, 1.25) |
| Hypertension | n (%) | 1008 (45.06) | 1274 (40.94) | 1245 (44.32) | 1462 (34.76) | 959 (39.64) |  |
|  | OR (95% CI) | Ref | 0.83**<br>(0.74, 0.94) | 0.80**<br>(0.71, 0.91) | 0.64***<br>(0.57, 0.72) | 0.66***<br>(0.58, 0.75) | 0.92***<br>(0.90, 0.94) |
| Antihypertensive<br>treatment | n (%) | 134 (6.03) | 560 (18.03) | 720 (25.63) | 1066 (25.28) | 815 (33.13) |  |
|  | OR (95% CI) | Ref | 3.22***<br>(2.63, 3.94) | 4.31***<br>(3.53, 5.27) | 5.37***<br>(4.42, 6.51) | 7.22***<br>(5.92, 8.79) | 1.32***<br>(1.28, 1.35) |
| Diabetes treatment | n (%) | 67 (2.98) | 109 (3.50) | 83 (2.96) | 176 (4.16) | 165 (6.70) |  |
|  | OR (95% CI) | Ref | 1.10 | 0.77 | 1.25 | 2.06*** | 1.15*** |
|  |  |  | (0.80, 1.50) | (0.56, 1.08) | (0.93, 1.67) | (1.54, 2.75) | (1.01, 1.21) |
| BMI, kg/m2 | mean ± SD | 25.7±4.5 | 25.7±4.7 | 26.2±5.3 | 27.1±6.0 | 27.7±6.1 |  |
|  | beta (95% CI) | Ref | 0.05<br>(-0.25, 0.35) | 0.51**<br>(0.20, 0.82) | 1.51***<br>(1.23, 1.80) | 1.99***<br>(1.67, 2.31) | 0.43***<br>(0.38, 0.49) |
| Total cholesterol | mean ± SD | 255.04±45.96 | 226±45 | 211±41 | 198±36 | 192±36 |  |
|  | beta (95% CI) | Ref | -28.27***<br>(-30.48, -26.06) | -45.11***<br>(-47.38, -42.85) | -54.79***<br>(-56.89, -52.69) | -64.16***<br>(-66.49, -61.82) | -11.46***<br>(-11.84, -11.07) |
| Total cholesterol/<br>HDL ratio |  | -- | 3.75±1.44 | 4.16±1.52 | 3.49±1.13 | 3.05±0.96 |  |
|  | beta (95% CI) | -- | Ref | 0.22***<br>(0.13, 0.31) | -0.39***<br>(-0.47, -0.31) | -0.89***<br>(-0.98, -0.80) | -0.28***<br>(-0.30, 0.26) |

<sup>a</sup>unadjusted descriptive statistics within each epoch and sex
<sup>b</sup>Age-adjusted differences compared to the reference epoch 1
<sup>c</sup>Trend estimated by regressing the baseline characteristic on time from each epoch-specific median date to the median date in epoch 1, adjusting for age. \*0.01<p<0.05, \*\*0.001<p<0.01, \*\*\*p<0.001

**Table 1B:** Baseline Characteristics and Incident Stroke Events in Each Epoch, in men.

| Baseline Characteristics |  | Men (n=12,574) |  |  |  |  |  |
| --- | --- | --- | --- | --- | --- | --- | --- |
|  |  | Epoch1 | Epoch 2 | Epoch 3 | Epoch 4 | Epoch 5 | Trend |
|  |  | 1962-67<br>n=1717 | 1971-76<br>n=2665 | 1987-91<br>n=2324 | 1998-2005<br>n=3405 | 2015-2019<br>n=2463 |  |
| Age, years | mean ± SD<sup>a</sup> | 57±8 | 55±12 | 58±13 | 55±13 | 62±12 | |
|  | beta (95% CI)<sup>b</sup> | Ref | -2.52***<br>(-3.25, -1.78) | 0.95*<br>(0.20, 1.70) | -2.21***<br>(-2.91, -1.51) | 5.16***<br>(4.41, 5.90) | 0.91***<br>(0.79, 1.04) |
| Age < 60 years | n (%) | 1096 (63.83) | 1732 (64.99) | 1331 (57.27) | 2301 (67.58) | 1036 (42.06) |  |
|  | OR (95% CI) | Ref | 0.95<br>(0.84, 1.08) | 1.32***<br>(1.16, 1.50) | 0.85**<br>(0.75, 0.96) | 2.43***<br>(2.14, 2.76) | 1.14***<br>(1.12, 1.17) |
| Current smoking | n (%) | 822 (52.73) | 924 (35.74) | 478 (20.63) | 456 (13.40) | 88 (4.45) |  |
|  | OR (95% CI) | Ref | 0.45***<br>(0.39, 0.51) | 0.23***<br>(0.20, 0.27) | 0.12***<br>(0.11, 0.14) | 0.04***<br>(0.03, 0.06) | 0.57***<br>(0.55, 0.59) |
| Prevalent CVD | n (%) | 237 (13.80) | 359 (13.47) | 400 (17.21) | 383 (11.25) | 215 (10.87) |  |
|  | OR (95% CI) | Ref | 0.98<br>(0.81, 1.18) | 0.99<br>(0.83, 1.20) | 0.71***<br>(0.59, 0.85) | 0.58***<br>(0.47, 0.71) | 0.89***<br>(0.87, 0.93) |
| AF | n (%) | 22 (1.28) | 46 (1.73) | 95 (4.09) | 158 (4.64) | 160 (8.09) |  |
|  | OR (95% CI) | Ref | 1.20<br>(0.71, 2.01) | 2.04<br>(1.27, 3.30) | 2.92***<br>(1.85, 4.62) | 5.26***<br>(3.34, 8.30) | 1.40***<br>(1.32, 1.49) |
| DM | n (%) | 63 (4.21) | 164 (6.48) | 170 (7.46) | 335 (10.17) | 295 (15.72) |  |
|  | OR (95% CI) | Ref | 1.65**<br>(1.22, 2.23) | 1.61**<br>(1.19, 2.17) | 2.72***<br>(2.06, 3.60) | 3.91***<br>(2.95, 5.19) | 1.28***<br>(1.23, 1.33) |
| Hypertension | n (%) | 721 (42.19) | 1114 (41.94) | 1137 (49.03) | 1357 (39.95) | 948 (48.64) |  |
|  | OR (95% CI) | Ref | 1.12<br>(0.98, 1.27) | 1.27**<br>(1.11, 1.45) | 1.01<br>(0.89, 1.14) | 1.17*<br>(1.02, 1.34) | 1.01<br>(0.99, 1.03) |
| Antihypertensive treatment | n (%) | 60 (3.52) | 335 (12.62) | 597 (25.78) | 909 (26.76) | 791 (39.99) |  |
|  | OR (95% CI) | Ref | 4.20***<br>(3.16, 5.59) | 8.78***<br>(6.65, 11.59) | 11.56***<br>(8.81, 15.17) | 17.58***<br>(13.34, 23.16) | 1.50***<br>(1.46, 1.55) |
| Diabetes treatment | n (%) | 57 (3.32) | 116 (4.36) | 107 (4.62) | 215 (6.32) | 220 (11.12) |  |
|  | OR (95% CI) | ref | 1.36<br>(0.98, 1.88) | 1.17<br>(0.84, 1.64) | 1.93***<br>(1.43, 2.60) | 3.20***<br>(2.37, 4.32) | 1.25***<br>(1.19, 1.31) |
| BMI, kg/m2 | mean ± SD | 26.3±3.4 | 26.9±3.5 | 27.6±3.9 | 28.5±4.6 | 29.2±4.9 | |
|  | beta (95% CI) | Ref | 0.54***<br>(0.28, 0.79) | 1.28***<br>(1.02, 1.54) | 2.11***<br>(1.87, 2.36) | 2.94***<br>(2.67, 3.22) | 0.57***<br>(0.53, 0.62) |
| Total cholesterol | mean ± SD | 238±42 | 217±39 | 205±38 | 194±37 | 174±37 | |
|  | beta (95% CI) | Ref | -22.28*** | -32.82*** | -45.10*** | -63.83*** | -11.09*** |
|  |  |  | (-24.61, -19.96) | (-35.21, -30.43) | (-47.33, -42.87) | (-66.33, -61.33) | (-11.50, -10.69) |
| Total cholesterol/<br>HDL ratio |  | --- | 5.26±1.88 | 5.15±1.65 | 4.50±1.41 | 3.68±1.21 |  |
|  | beta (95% CI) | -- | Ref | -0.02<br>(-0.13, 0.09) | -0.69***<br>(-0.79, -0.60) | -1.48***<br>(-1.60, -1.37) | -0.40***<br>(-0.42, -0.37) |

<sup>a</sup>unadjusted descriptive statistics within each epoch and sex
<sup>b</sup>Age-adjusted differences compared to the reference epoch 1
<sup>c</sup>Trend estimated by regressing the baseline characteristic on time from each epoch-specific median date to the median date in epoch 1, adjusting for age. \*0.01<p<0.05, \*\*0.001<p<0.01, \*\*\*p<0.001

**Table 2:** Interaction between epoch or trends by sex on baseline characteristics.

|  | Epoch<br>(p value) | Trends<br>(p value) |
| --- | --- | --- |
| Age, years | <0.001 | 0.06 |
| Age < 60 years | 0.88 | 0.62 |
| Current smoking | <0.001 | <0.001 |
| Prevalent CVD | 0.50 | 0.81 |
| AF | 0.01 | 0.03 |
| DM | 0.06 | 0.04 |
| Hypertension | <0.001 | <0.001 |
| Antihypertensive treatment | <0.001 | <0.001 |
| Diabetes treatment | 0.19 | 0.02 |
| BMI, kg/m<sup>2</sup> | <0.001 | <0.001 |
| Total cholesterol | <0.001 | 0.20 |
| Total cholesterol/ HDL ratio | <0.001 | <0.001 |

### Incident stroke in men compared to women by epoch

Although men had a higher stroke risk than women across all epochs, the differences reached statistical significance only in epoch 2 (HR 1.41, 95% CI [1.08, 1.84], p <0.05) and epoch 4 (HR 1.46, 95% CI [1.11, 1.91], p <0.01). (Table 3A).

**Table 3A:** Incident stroke in men compared to women, and for all participants within epochs.

|  | All participants |  |
| --- | --- | --- |
| epoch | Stroke cases/total observations | Hazard ratio (95% CI) |
| Epoch 1 (1962-69) | 178/3966 | 1.24 (0.92, 1.67) |
| Epoch 2 (1971-76) | 227/5786 | 1.41 (1.08, 1.84)* |
| Epoch 3 (1987-91) | 199/5137 | 1.22 (0.93, 1.62) |
| Epoch 4 (1998-2005) | 207/7642 | 1.46 (1.11, 1.91)** |
| Epoch 5 (2015-2021) | 119/5534 | 1.30 (0.91, 1.86) |

Reference is women. Models are adjusted for age. \* $0.01 < p < 0.05$ , \*\* $0.001 < p < 0.01$ ,
\*\*\* $p < 0.001$

**Table 3B:** Incident stroke in men compared to women for participants < 60 years old, within epochs.

|  | Participants < 60 years |  |
| --- | --- | --- |
| epoch | Stroke cases/total observations | Hazard ratio (95% CI) |
| Epoch 1 (1962-69) | 53/2477 | 1.39 (0.81, 2.38) |
| Epoch 2 (1971-76) | 50/3511 | 1.02 (0.58, 1.77) |
| Epoch 3 (1987-91) | 27/2840 | 1.92 (0.88, 4.18) |
| Epoch 4 (1998-2005) | 46/5068 | 2.13 (1.17, 3.87)* |
| Epoch 5 (2015-2021) | 11/2320 | 3.45 (0.92, 13.02) |

Reference is women. Models are adjusted for age. \* $0.01 < p < 0.05$ , \*\* $0.001 < p < 0.01$ ,
\*\*\* $p < 0.001$

### Trends in Stroke incidence

Relative to the first epoch, stroke incidence in all participants declined by 24%, 42%, 53% and 64% in subsequent epochs (p<0.01 for all epochs, Table 4A). In women, stroke incidence declined by 31%, 46%, 59%, 68% in subsequent epochs (p<0.01 for all epochs) with an estimated decline of 19% per decade (p<0.001). In men, stroke incidence declined by 17%, 40%, 47%, 62% in successive epochs in men (p<0.01 for epochs 3-5) with an estimated decline of 16% per decade (p<0.001). There was no significant interaction between sex and epoch or sex and trend.

**Table 4A:** Temporal Trends in 10-Year incidence of stroke, overall, and stratified by Sex.

|  | No.<br>Cases | No.<br>Observations | Epoch 1 | Epoch 2<br>HR (95% CI) | Epoch 3<br>HR (95% CI) | Epoch 4<br>HR (95% CI) | Epoch 5<br>HR (95% CI) | Hazard Ratio<br>Trend |
| --- | --- | --- | --- | --- | --- | --- | --- | --- |
| All<br>participants | 930 | 28,065 | REF | 0.76<br>(0.62, 0.92) | 0.58<br>(0.47, 0.71) | 0.47<br>(0.38, 0.58) | 0.36<br>(0.28, 0.45) | 0.82<br>(0.79, 0.86) ** |
| Women | 499 | 15,491 | REF | 0.69<br>(0.52, 0.91) | 0.54<br>(0.40, 0.71) | 0.41<br>(0.30, 0.54) | 0.32<br>(0.23, 0.45) | 0.81<br>(0.76, 0.86) ** |
| Men | 431 | 12,574 | REF | 0.83<br>(0.62, 1.11) | 0.60<br>(0.44, 0.82) | 0.53<br>(0.40, 0.72) | 0.38<br>(0.27, 0.54) | 0.84<br>(0.79, 0.89) ** |

<sup>#</sup>Baseline FHS examinations occurred within 5 epochs: Epoch 1, 1962 to 1967; Epoch 2, 1971 to 1976; Epoch 3, 1987 to 1991; Epoch 4, 1998 to 2005 and epoch 5, 2015 to 2021. Ten-year hazards ratios (the incidence of stroke during each epoch relative to the incidence during the first epoch), adjusted for age at baseline. \* p for trend < 0.01, \*\* p for trend < 0.001

**Table 4B:** Temporal Trends in 10-Year incidence of stroke, in those < 60 years of age, and stratified by Sex.

|  | No.<br>Cases | No.<br>Observations | Epoch 1 | Epoch 2<br>HR (95% CI) | Epoch 3<br>HR (95% CI) | Epoch 4<br>HR (95% CI) | Epoch 5<br>HR (95% CI) | Hazard Ratio<br>Trend |
| --- | --- | --- | --- | --- | --- | --- | --- | --- |
| < 60 years | 187 | 16,216 | REF | 0.87<br>(0.59, 1.28) | 0.54<br>(0.34, 0.86) | 0.56<br>(0.38, 0.83) | 0.62<br>(0.32, 1.22) | 0.86<br>(0.79, 0.95) * |
| Women | 82 | 8720 | REF | 1.00<br>(0.57, 1.73) | 0.44<br>(0.21, 0.90) | 0.43<br>(0.23, 0.80) | 0.38<br>(0.11, 1.29) | 0.78<br>(0.67, 0.90) ** |
| Men | 105 | 7496 | REF | 0.74<br>(0.43, 1.28) | 0.62<br>(0.34, 1.13) | 0.67<br>(0.40, 1.14) | 0.83<br>(0.37, 1.88) | 0.94<br>(0.82, 1.06) |

#Baseline FHS examinations occurred within 5 epochs: Epoch 1, 1962 to 1967; Epoch 2, 1971 to 1976; Epoch 3, 1987 to 1991; Epoch 4, 1998 to 2005 and epoch 5, 2015 to 2021. Ten-year hazards ratios (the incidence of stroke during each epoch relative to the incidence during the first epoch), adjusted for age at baseline.
\* p for trend < 0.01, \*\* p for trend < 0.001

### Secondary analysis

In analyses restricting the sample to participants aged < 60 years, we observed higher stroke risk in men compared to women over all five epochs but reaching statistical significance only in epoch 4 (HR 2.13, 95% CI 1.17, 3.87, p <0.05, Table 3B). There was no significant interaction between sex and epoch, but we observed a significant interaction between sex and trend (p=0.05). In women < 60 years, relative to the first epoch, stroke incidence showed no decline in the second epoch and an increasing decline by 56%, 57% and 62% for subsequent epochs (p<0.01 for epochs 3 and 4), with an estimated decline of 22% per epoch (p<0.001) (Table 4B). In contrast, in men < 60 years, stroke incidence showed more variability, with overall decreasing degree of decline over time: by 26%, 38%, 33% and 17% in subsequent epochs (all with p>0.05), with an estimated decline of 6% per epoch (p>0.05) (Table 4B).

## Discussion

In our cohort of community dwelling adults with up to seven decades of prospective follow-up, we observed sex-specific differences in stroke incidence and secular trends in stroke, which were dependent on age. Stroke risk was higher in men across all epochs, and while there was a decline in stroke incidence in both men and women, the decline observed was to a lesser degree and showed a decreasing trend in men (i.e. less decline over time).

Among participants aged < 60 years, stroke risk was higher in men with a stronger difference than that noted in the overall sample, and a significant decline in stroke incidence occurred only in women. Evaluation of trends in vascular risk factors showed that smoking and prevalent CVD declined, while atrial fibrillation, diabetes, treatments for diabetes and hypertension increased in both sexes. Importantly, hypertension trends declined only in women.

This study provides updated data in stroke incidence and sex specific trends in stroke in a large prospective cohort with the longest follow up available. It adds to existing knowledge about the decline in stroke incidence previously observed, showing a continued decline beyond previously reported periods in US and non-US studies.^13,42,43^ Further, while previous studies noted a decline in stroke risk only in older people (> 65 years^13,43^ and > 80 years^42^), we observed a 64% reduction in risk for stroke in the last epoch (2015 -2021) compared to the first (1962-67) in all participants and 38% in those < 60 years, and similar decline in trends in both groups. However, we observed sex specific differences such that the decline in stroke incidence trends in those < 60 years was driven by reduced trends only in women. Previous studies^13,42^ have not observed any sex differences in decline of stroke, though these studies were restricted to older people. Although the reason for the sex difference in individuals < 60 years may be multifactorial, we observed a declining trend for hypertension in women over time which was not observed in men. Hypertension is among the strongest if not the strongest risk factor for stroke, thus it would not be surprising that it accounts for the sex specific differences observed. In exploring the trends in other modifiable stroke risk factors, our study showed similar trends in both sexes. We observed encouraging decline overtime in certain risk factors like smoking and prevalent cardiovascular disease in both sexes but increasing trends in atrial fibrillation and diabetes in both sexes. Prior studies from the FHS^37,44^ showed that over 50 years of observation (1958-2007),^44^ the prevalence and incidence of atrial fibrillation increased in both sexes, and in men between 1950 -2004.^37^ The current study expands on this earlier finding showing that more than a decade later, the prevalence of atrial fibrillation is increasing. Enhanced surveillance for atrial fibrillation and improvement in diagnostic detection methods may account for the rising prevalence noted previously. Although the trends for obesity declined in both sexes over time, we observed that compared to the first epoch, the prevalence of obesity increased in the last two epochs in both sexes. This observation underscores the rising prevalence of obesity.^45^ The increased trend in diabetes highlights the need to target metabolic disorders in public health interventions, possibly linked to the increasing BMI in later epochs in both sexes,which reflect the obesity epidemic in the US and globally.^46^

Despite increasing trends in use of hypertension treatment over time in both sexes, only women had reduced odds of and trends for having hypertension while men had increased long-term risk for hypertension. Hypertension remains a major target for stroke prevention, yet it remains unrecognized and undertreated in large segments of the population.^3^ Our results bring attention to the need for further preventive public health efforts, particularly in men.

Among the strengths of our study are the long-term follow-up with high participant retention and rigorous surveillance for incident stroke events. Stroke diagnosis is adjudicated by an expert panel, enhancing diagnostic accuracy. The large sample size and extended duration of follow-up, the longest in epidemiological research, enabled robust evaluation of temporal trends in stroke incidence and associated risk factors across decades. In addition, the inclusion of participants < 60 years allowed for assessment of stroke trends in younger individuals. Several limitations should also be acknowledged. The cohort is composed predominantly of individuals of White race, which limits generalizability of findings to more diverse populations.

## Conclusion

Stroke incidence and secular trends in stroke incidence continue to decline across all ages beyond earlier observed periods. However, among those < 60 years, significant decline was observed only in women. Trends in modifiable vascular risk factors differed by sex and may partially explain the observed sex differences in stroke incidence, particularly with respect to hypertension. Atrial fibrillation and diabetes have increased over time in both sexes, while hypertension declined only in women.

## Clinical implications

Despite continued declines in stroke incidence, the rising prevalence of key risk factors, especially atrial fibrillation and diabetes, underscores the need for enhanced surveillance for these stroke risk factors to reduce the burden of stroke. Modifiable risk factors remain as essential treatment targets to reduce stroke risk. Additionally, the increased risk of hypertension over time in men, with reduced trends observed only in women <60 years suggests higher risk in younger men. Ongoing public health interventions are needed to increase awareness and treatment of modifiable risk factors.

## CRediT authorship contribution statement

Oluchi Ekenze: Writing – original draft, Writing – review & editing, Conceptualization. Mathew Scott: Writing – review & editing, Writing – original draft, Formal analysis, Data curation. Dibya Himali: Writing – review & editing, Formal analysis, Data curation.

Vasileios-Arsenios Lioutas: Writing – review & editing. Sudha Seshadri: Writing – review & editing, Fund acquisition. Virginia J. Howard: Writing – review & editing. Myriam Fornage: Writing – review & editing. Hugo J. Aparicio: Writing – review & editing, Fund acquisition. Alexa S. Beiser: Writing – review & editing, Writing – original draft, Formal analysis, Data curation. Jose R. Romero: Writing – review & editing, Writing – original draft, Data curation, Conceptualization, Fund acquisition.

## Data Availability

Requests to access de-identified data from the Framingham Heart Study can be made at https://www.framinghamheartstudy.org/fhs-for-researchers

## References

1. Vyas MV, Silver FL, Austin PC, Yu AYX, Pequeno P, Fang J, Laupacis A, Kapral MK. Stroke Incidence by Sex Across the Lifespan. Stroke. 2021;52:447–451. doi: 10.1161/STROKEAHA.120.032898

2. Cheng Y, Lin Y, Shi H, Cheng M, Zhang B, Liu X, Shi C, Wang Y, Xia C, Xie W. Projections of the Stroke Burden at the Global, Regional, and National Levels up to 2050 Based on the Global Burden of Disease Study 2021. J Am Heart Assoc. 2024;13:e036142. doi: 10.1161/JAHA.124.036142

3. Palaniappan LP, Allen NB, Almarzooq ZI, Anderson CAM, Arora P, Avery CL, Baker-Smith CM, Bansal N, Currie ME, Earlie RS, et al. 2026 Heart Disease and Stroke Statistics: A Report of US and Global Data From the American Heart Association. Circulation. 2026;153:e275–e906. doi: 10.1161/CIR.0000000000001412

4. Appelros P, Stegmayr B, Terent A. Sex differences in stroke epidemiology: a systematic review. Stroke. 2009;40:1082–1090. doi: 10.1161/STROKEAHA.108.540781

5. Howard VJ, Madsen TE, Kleindorfer DO, Judd SE, Rhodes JD, Soliman EZ, Kissela BM, Safford MM, Moy CS, McClure LA, et al. Sex and Race Differences in the Association of Incident Ischemic Stroke With Risk Factors. JAMA Neurol. 2019;76:179–186. doi: 10.1001/jamaneurol.2018.3862

6. Petrea RE, Beiser AS, Seshadri S, Kelly-Hayes M, Kase CS, Wolf PA. Gender differences in stroke incidence and poststroke disability in the Framingham heart study. Stroke. 2009;40:1032–1037. doi: 10.1161/STROKEAHA.108.542894

7. Islam MS, Anderson CS, Hankey GJ, Hardie K, Carter K, Broadhurst R, Jamrozik K. Trends in incidence and outcome of stroke in Perth, Western Australia during 1989 to 2001: the Perth Community Stroke Study. Stroke. 2008;39:776–782. doi: 10.1161/STROKEAHA.107.493643

8. Madsen TE, Khoury J, Alwell K, Moomaw CJ, Rademacher E, Flaherty ML, Woo D, Mackey J, De Los Rios La Rosa F, Martini S, et al. Sex-specific stroke incidence over time in the Greater Cincinnati/Northern Kentucky Stroke Study. Neurology. 2017;89:990–996. doi: 10.1212/WNL.0000000000004325

9. Global, regional, and national burden of stroke and its risk factors, 1990-2021: a systematic analysis for the Global Burden of Disease Study 2021. Lancet Neurol. 2024;23:973–1003. doi: 10.1016/s1474-4422(24)00369-7

10. He Q, Wang W, Zhang Y, Xiong Y, Tao C, Ma L, Ma J, You C, Wang C. Global, Regional, and National Burden of Stroke, 1990-2021: A Systematic Analysis for Global Burden of Disease 2021. Stroke. 2024;55:2815–2824. doi: 10.1161/STROKEAHA.124.048033

11. Terent A. Trends in stroke incidence and 10-year survival in Soderhamn, Sweden, 1975-2001. Stroke. 2003;34:1353–1358. doi: 10.1161/01.STR.0000074038.71700.1C

12. Benatru I, Rouaud O, Durier J, Contegal F, Couvreur G, Bejot Y, Osseby GV, Ben Salem D, Ricolfi F, Moreau T, et al. Stable stroke incidence rates but improved case-fatality in Dijon, France, from 1985 to 2004. Stroke. 2006;37:1674–1679. doi: 10.1161/01.STR.0000226979.56456.a8

13. Koton S, Sang Y, Schneider ALC, Rosamond WD, Gottesman RF, Coresh J. Trends in Stroke Incidence Rates in Older US Adults: An Update From the Atherosclerosis Risk in Communities (ARIC) Cohort Study. JAMA Neurol. 2020;77:109–113. doi: 10.1001/jamaneurol.2019.3258

14. Lisabeth LD, Brown DL, Zahuranec DB, Kim S, Lim J, Kerber KA, Meurer WJ, Case E, Smith MA, Campbell MS, et al. Temporal Trends in Ischemic Stroke Rates by Ethnicity, Sex, and Age (2000-2017): The Brain Attack Surveillance in Corpus Christi Project. *Neurology*. 2021;97:e2164–e2172. doi: 10.1212/WNL.0000000000012877

15. Madsen TE, Khoury JC, Leppert M, Alwell K, Moomaw CJ, Sucharew H, Woo D, Ferioli S, Martini S, Adeoye O, et al. Temporal Trends in Stroke Incidence Over Time by Sex and Age in the GCNKSS. Stroke. 2020;51:1070–1076. doi: 10.1161/STROKEAHA.120.028910

16. Seshadri S, Beiser A, Kelly-Hayes M, Kase CS, Au R, Kannel WB, Wolf PA. The lifetime risk of stroke: estimates from the Framingham Study. Stroke. 2006;37:345–350. doi: 10.1161/01.STR.0000199613.38911.b2

17. Collaborators GBDLRoS, Feigin VL, Nguyen G, Cercy K, Johnson CO, Alam T, Parmar PG, Abajobir AA, Abate KH, Abd-Allah F, et al. Global, Regional, and Country-Specific Lifetime Risks of Stroke, 1990 and 2016. N Engl J Med. 2018;379:2429–2437. doi: 10.1056/NEJMoa1804492

18. O’Donnell MJ, Chin SL, Rangarajan S, Xavier D, Liu L, Zhang H, Rao-Melacini P, Zhang X, Pais P, Agapay S, et al. Global and regional effects of potentially modifiable risk factors associated with acute stroke in 32 countries (INTERSTROKE): a case-control study. Lancet. 2016;388:761–775. doi: 10.1016/S0140-6736(16)30506-2

19. Madsen TE, Howard G, Kleindorfer DO, Furie KL, Oparil S, Manson JE, Liu S, Howard VJ. Sex Differences in Hypertension and Stroke Risk in the REGARDS Study: A Longitudinal Cohort Study. Hypertension. 2019;74:749–755. doi: 10.1161/HYPERTENSIONAHA.119.12729

20. Peters SAE, Carcel C, Millett ERC, Woodward M. Sex differences in the association between major risk factors and the risk of stroke in the UK Biobank cohort study. Neurology. 2020;95:e2715–e2726. doi: 10.1212/WNL.0000000000010982

21. Fang MC, Singer DE, Chang Y, Hylek EM, Henault LE, Jensvold NG, Go AS. Gender differences in the risk of ischemic stroke and peripheral embolism in atrial fibrillation: the AnTicoagulation and Risk factors In Atrial fibrillation (ATRIA) study. Circulation. 2005;112:1687–1691. doi: 10.1161/CIRCULATIONAHA.105.553438

22. Wang TJ, Massaro JM, Levy D, Vasan RS, Wolf PA, D’Agostino RB, Larson MG, Kannel WB, Benjamin EJ. A risk score for predicting stroke or death in individuals with new-onset atrial fibrillation in the community: the Framingham Heart Study. JAMA. 2003;290:1049–1056. doi: 10.1001/jama.290.8.1049

23. Dagres N, Nieuwlaat R, Vardas PE, Andresen D, Levy S, Cobbe S, Kremastinos DT, Breithardt G, Cokkinos DV, Crijns HJ. Gender-related differences in presentation, treatment, and outcome of patients with atrial fibrillation in Europe: a report from the Euro Heart Survey on Atrial Fibrillation. J Am Coll Cardiol. 2007;49:572–577. doi: 10.1016/j.jacc.2006.10.047

24. Giralt D, Domingues-Montanari S, Mendioroz M, Ortega L, Maisterra O, Perea-Gainza M, Delgado P, Rosell A, Montaner J. The gender gap in stroke: a meta-analysis. Acta Neurol Scand. 2012;125:83–90. doi: 10.1111/j.1600-0404.2011.01514.x

25. Dufouil C, Beiser A, McLure LA, Wolf PA, Tzourio C, Howard VJ, Westwood AJ, Himali JJ, Sullivan L, Aparicio HJ, et al. Revised Framingham Stroke Risk Profile to Reflect Temporal Trends. Circulation. 2017;135:1145–1159. doi: 10.1161/CIRCULATIONAHA.115.021275

26. Marzona I, Proietti M, Farcomeni A, Romiti GF, Romanazzi I, Raparelli V, Basili S, Lip GYH, Nobili A, Roncaglioni MC. Sex differences in stroke and major adverse clinical events in patients with atrial fibrillation: A systematic review and meta-analysis of 993,600 patients. Int J Cardiol. 2018;269:182–191. doi: 10.1016/j.ijcard.2018.07.044

27. Maeda T, Nishi T, Funakoshi S, Tada K, Tsuji M, Satoh A, Kawazoe M, Yoshimura C, Arima H. Risk of Stroke in Atrial Fibrillation According to Sex in Patients Aged Younger Than 75 Years: A Large-Scale, Observational Study Using Real-World Data. Heart Lung Circ. 2021;30:963–970. doi: 10.1016/j.hlc.2020.11.012

28. Cameron AJ, Magliano DJ, Soderberg S. A systematic review of the impact of including both waist and hip circumference in risk models for cardiovascular diseases, diabetes and mortality. Obes Rev. 2013;14:86–94. doi: 10.1111/j.1467-789X.2012.01051.x

29. Rodriguez-Campello A, Jimenez-Conde J, Ois A, Cuadrado-Godia E, Giralt-Steinhauer E, Vivanco RM, Soriano-Tarraga C, Subirana I, Munoz D, Gomez-Gonzalez A, et al. Sex-related differences in abdominal obesity impact on ischemic stroke risk. Eur J Neurol. 2017;24:397–403. doi: 10.1111/ene.13216

30. Towfighi A, Zheng L, Ovbiagele B. Weight of the obesity epidemic: rising stroke rates among middle-aged women in the United States. Stroke. 2010;41:1371–1375. doi: 10.1161/STROKEAHA.109.577510

31. Zhao W, Katzmarzyk PT, Horswell R, Wang Y, Johnson J, Hu G. Sex differences in the risk of stroke and HbA(1c) among diabetic patients. Diabetologia. 2014;57:918–926. doi: 10.1007/s00125-014-3190-3

32. Peters SA, Huxley RR, Woodward M. Diabetes as a risk factor for stroke in women compared with men: a systematic review and meta-analysis of 64 cohorts, including 775,385 individuals and 12,539 strokes. Lancet. 2014;383:1973–1980. doi: 10.1016/S0140-6736(14)60040-4

33. Carcel C, Sandset EC, Ali M, Allende Echanez MI, Mosconi MG, de Souza AC, Dalli LL, Venturelli PM, Sakamoto Y, Nasreldein A, et al. Addressing sex and gender differences in stroke risk and management: A scientific statement from the World Stroke Organization. Int J Stroke. 2026;21:303–323. doi: 10.1177/17474930251393009

34. Madsen TE, Howard VJ, Jimenez M, Rexrode KM, Acelajado MC, Kleindorfer D, Chaturvedi S. Impact of Conventional Stroke Risk Factors on Stroke in Women: An Update. Stroke. 2018;49:536–542. doi: 10.1161/STROKEAHA.117.018418

35. Tsao CW, Vasan RS. Cohort Profile: The Framingham Heart Study (FHS): overview of milestones in cardiovascular epidemiology. Int J Epidemiol. 2015;44:1800–1813. doi: 10.1093/ije/dyv337

36. Aparicio HJ, Himali JJ, Satizabal CL, Pase MP, Romero JR, Kase CS, Beiser AS, Seshadri S. Temporal Trends in Ischemic Stroke Incidence in Younger Adults in the Framingham Study. Stroke. 2019;50:1558–1560. doi: 10.1161/STROKEAHA.119.025171

37. Carandang R, Seshadri S, Beiser A, Kelly-Hayes M, Kase CS, Kannel WB, Wolf PA. Trends in incidence, lifetime risk, severity, and 30-day mortality of stroke over the past 50 years. JAMA. 2006;296:2939–2946. doi: 10.1001/jama.296.24.2939

38. Chobanian AV, Bakris GL, Black HR, Cushman WC, Green LA, Izzo JL, Jr., Jones DW, Materson BJ, Oparil S, Wright JT, Jr., et al. The Seventh Report of the Joint National Committee on Prevention, Detection, Evaluation, and Treatment of High Blood Pressure: the JNC 7 report. JAMA. 2003;289:2560–2572. doi: 10.1001/jama.289.19.2560

39. Ekenze O, Pinheiro A, Beiser AS, Lioutas VA, Aparicio HJ, Benjamin EJ, Vasan RS, DeCarli C, Seshadri S, Demissie S, et al. Prevalent Cardiovascular Disease and Atrial Fibrillation in Relation to Cerebral Small Vessel Disease Burden. Brain Sci. 2025;15. doi: 10.3390/brainsci15080813

40. Lin DY, Wei LJ. The Robust Inference for the Cox Proportional Hazards Model. Journal of the American Statistical Association. 1989;84:1074–1078. doi: 10.1080/01621459.1989.10478874

41. Koellhoffer EC, McCullough LD. The effects of estrogen in ischemic stroke. Transl Stroke Res. 2013;4:390–401. doi: 10.1007/s12975-012-0230-5

42. Cerasuolo JO, Cipriano LE, Sposato LA, Kapral MK, Fang J, Gill SS, Hackam DG, Hachinski V. Population-based stroke and dementia incidence trends: Age and sex variations. Alzheimers Dement. 2017;13:1081–1088. doi: 10.1016/j.jalz.2017.02.010

43. Aked J, Delavaran H, Norrving B, Lindgren A. Temporal Trends of Stroke Epidemiology in Southern Sweden: A Population-Based Study on Stroke Incidence and Early Case-Fatality. Neuroepidemiology. 2018;50:174–182. doi: 10.1159/000487948

44. Schnabel RB, Yin X, Gona P, Larson MG, Beiser AS, McManus DD, Newton-Cheh C, Lubitz SA, Magnani JW, Ellinor PT, et al. 50 year trends in atrial fibrillation prevalence, incidence, risk factors, and mortality in the Framingham Heart Study: a cohort study. Lancet. 2015;386:154–162. doi: 10.1016/S0140-6736(14)61774-8

45. Ng M, Dai X, Cogen RM, Abdelmasseh M, Abdollahi A, Abdullahi A, Aboagye RG, Abukhadijah HJ, Adeyeoluwa TE, Afolabi AA, et al. National-level and state-level prevalence of overweight and obesity among children, adolescents, and adults in the USA, 1990&#x2013;2021, and forecasts up to 2050. The Lancet. 2024;404:2278–2298. doi: 10.1016/S0140-6736(24)01548-4

46. Martin SS, Aday AW, Almarzooq ZI, Anderson CAM, Arora P, Avery CL, Baker-Smith CM, Barone Gibbs B, Beaton AZ, Boehme AK, et al. 2024 Heart Disease and Stroke Statistics: A Report of US and Global Data From the American Heart Association. Circulation. 2024;149:e347–e913. doi: 10.1161/CIR.0000000000001209

